# Early-onset diabetes in Africa: A mini-review of the current genetic profile

**DOI:** 10.1101/2023.08.20.23294330

**Authors:** Samuel Mawuli Adadey, Joy Afua Mensah, Kojo Sekyi Acquah, Abugri James, Richard Osei-Yeboah

## Abstract

Early-onset diabetes is poorly diagnosed partly due to its heterogeneity and variable presentations. Although several genes have been associated with the disease, these genes are not well studied in Africa. We sought to identify the major neonatal, early childhood, juvenile, or early-onset diabetes genes in Africa; and evaluate the available molecular methods used for investigating these gene variants. A literature search was conducted on PubMed, Scopus, Africa-Wide Information, and Web of Science databases. The retrieved records were screened and analyzed to identify genetic variants associated with early-onset diabetes. Although 319 records were retrieved, 32 were considered for the current review. Most of these records (22/32) were from North Africa. The disease condition was genetically heterogenous with most cases possessing unique gene variants. We identified 22 genes associated with early-onset diabetes, 9 of which had variants (n=19) classified as pathogenic or likely pathogenic (PLP). Among the PLP variants, *IER3IP1*: p.(Leu78Pro) was the variant with the highest number of cases. There was limited data from West Africa, hence the contribution of genetic variability to early-onset diabetes in Africa could not be comprehensively evaluated. It is worth mentioning that most studies were focused on natural products as antidiabetics and only a few studies reported on the genetics of the disease. *ABCC8* and *KCNJ11* were implicated as major contributors to early-onset diabetes gene networks. Gene ontology analysis of the network associated ion channels, impaired glucose tolerance, and decreased insulin secretions to the disease. Our review highlights 9 genes from which PLP variants have been identified and can be considered for the development of an African diagnostic panel. There is a gap in early-onset diabetes genetic research from sub-Saharan Africa which is much needed to develop a comprehensive, efficient, and cost-effective genetic panel that will be useful in clinical practice on the continent and among the African diasporas.

## Introduction

Early-onset diabetes consists of a spectrum of diseases such as maturity-onset diabetes of the young (MODY), juvenile diabetes, neonatal diabetes mellitus, and rare diabetes-associated syndromic disease. These early-onset diabetic conditions may be associated with single genes and therefore characterized as monogenic diabetes [1]. Globally, about 500,000 children are diagnosed of Type 1 Diabetes Mellitus (T1DM) [2] with an estimated 3% annual increase [3]. Most of the T1DM cases were from Europe and North America [2]. A prevalence of 2.5% of monogenic diabetes was reported from United Kingdom pediatric clinics [4]. MODY has been estimated to account for 1-5% of diabetes cases [5] and advances in sequencing technologies has made it possible for 14 MODY genes to be identified [6, 7]. The contribution of these genes and other early-onset diabetes associated genes in Africa diabetes remains unknown. Hence, we hereby, present a literature review on the current genetic profile of early-onset diabetes in Africa.

Early-onset diabetes remains a challenge for affected individuals and their families and has an impact on quality of life. A diagnosis of diabetes generally influences a change in a patient’s life style, psychological and general well-being which is often accompanied by stress [8]. About 40% of diabetes patients have psychological problems which reduces their quality of life [9].Patients with severe cases are faced with a lifelong injection of insulin for patients with insulin dependent diabetes (T1DM) and daily pills for patients with insulin independent diabetes (Type 2 Diabetes Mellitus (T2DM)) [10]. Studies on insulin use and quality of life has shown that insulin use in MODY and T1DM cases corelates with poor quality of life [8, 11]. This observation may be attributed to the switch from pills to daily injections for some MODY cases and the lifelong dependency on insulin injections for T1DM which is not convenient [8]. It has also been reported that the affected individuals struggle with not having the freedom to eat or drink what they like and in some cases, they have a negative view of the future [8]. It is worth mentioning that there is a mix of the impact of early-onset diabetes on the education of children living with the condition [12]. Availability of effective management and treatment options is a crucial factor where the education of children in resource rich settings is often not affected, and the converse is true in resource poor settings.

The classical diagnosis of early-onset diabetes may be failing due to issues such as low penetrance observed in some MODY cases, several sub-classifications of the disease, and unusual clinical presentations of the disease [13-15]. Attempts to address these challenges have led to a global expansion of diagnostics and this was necessitated by an increasing public health concern [16]. Improvements in gene sequencing technologies and accessibility of genetic testing to patients has contributed to early diagnosis and improved treatment outcomes in patients [7, 14]. However, only a few of the early-onset diabetes cases are effectively diagnosed especially in developing countries [2]. Design and development of population specific genetic diagnostic panels can enhance screening for early-onset diabetes in Africa. We therefore sought to review the genes associated with early-onset diabetes in Africa to identify potential genes that can be considered in the design of a genetic panel.

## Method

Here, we reviewed the literature to investigate the major early-onset diabetes genes reported from patients of African ancestry. The protocol for this systematic review was registered with PROSPERO [17] (ID#: CRD42022324696). The review was conducted using Covidence, a systematic review software which allows multiple reviewers to work through the steps of a systematic review efficiently and effectively [18].

### Search strategy and screening

The keywords, early-onset, neonatal, juvenile, diabetes, genes, genetics, genomics, and Africa were used to develop the search term below. [((((((Early-onset) OR (Neonatal)) OR (Juvenile)) OR (childhood)) AND (((Genetics) OR (genes)) OR (genomics))) AND (Diabetes)) AND (Africa OR Algeria OR Angola OR Benin OR Botswana OR “Burkina Faso” OR Burundi OR Cameroon OR “Cape Verde” OR “Central African Republic” OR Chad OR Comoros OR Djibouti OR “DR Congo” OR “Democratic Republic of Congo”, OR Congo OR Egypt OR “Equatorial Guinea” OR Guinea OR Eritrea OR Eswatini OR Ethiopia OR Gabon OR Gambia OR Ghana OR Guinea OR (Guinea Bissau) OR “Ivory Coast” OR “Côte d’Ivoire” OR Kenya OR Lesotho OR Liberia OR Libya OR Madagascar OR Malawi OR Mali OR Mauritania OR Mauritius OR Morocco OR Mozambique OR Namibia OR Niger OR Nigeria OR Rwanda OR “Sao Tome And Principe” OR Senegal OR Seychelles OR Sierra Leone OR Somalia OR “South Africa” OR “South Sudan” OR Sudan OR Tanzania OR Togo OR Tunisia OR Uganda OR Zambia OR Zimbabwe)].

A literature search was conducted on PubMed, Scopus, Africa-Wide Information, and Web of Science databases from 31^st^ July to 20^th^ December 2022 by two independent reviewers. A total of 319 records were retrieved from the databases. Two reviewers screened the retrieved records using the title and abstract, and 32 publications were retained for data extraction (Figure 1). The inclusion and exclusion criteria below were used to screen the retrieved records.

**Figure 1.**
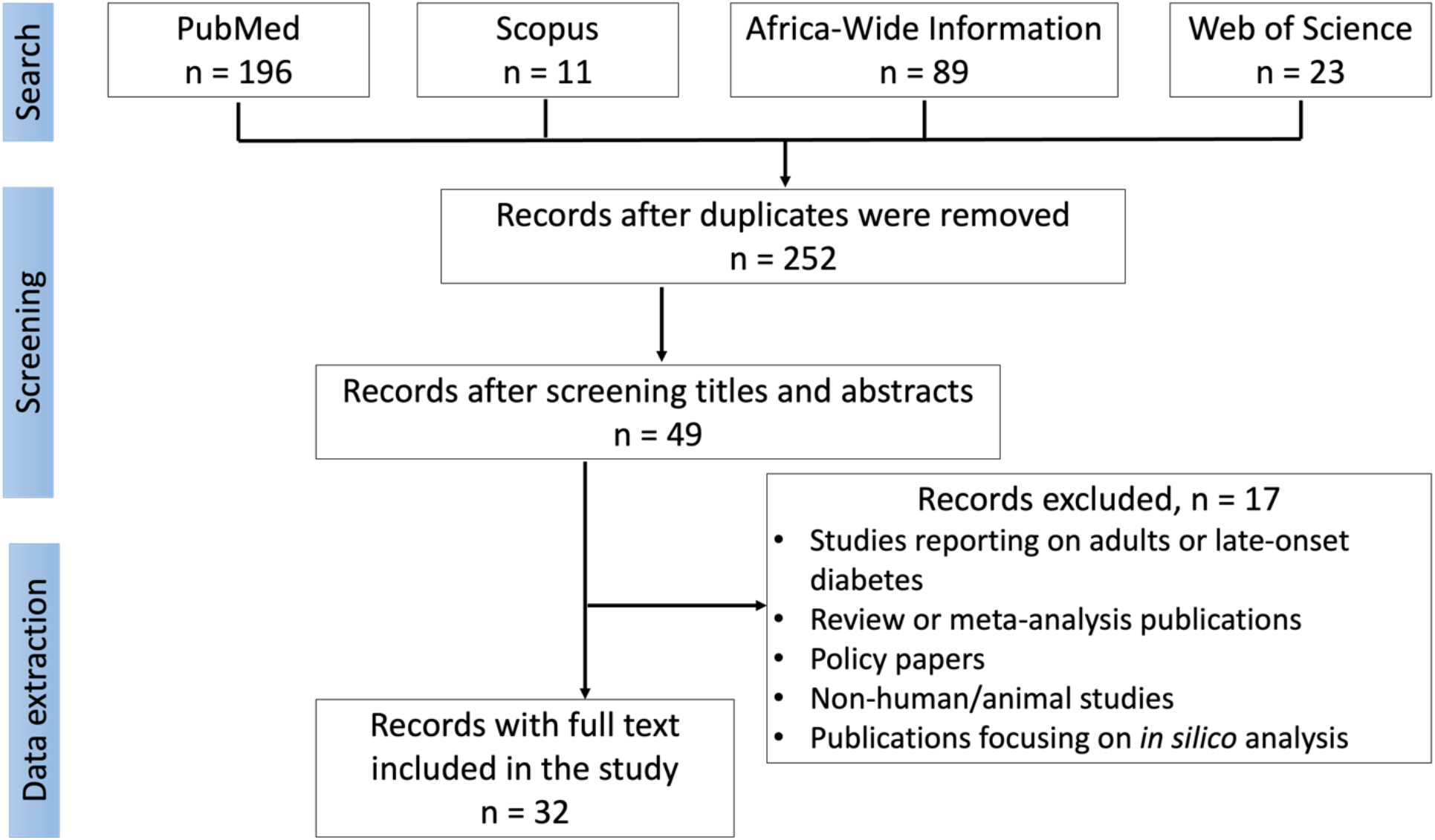
: A flow diagram of the search and screening strategy

Original research studies which met the following inclusion criteria were considered:

1. neonatal, early childhood, juvenile, or early-onset diabetes
2. studies reporting on gene variants associated with diabetics
3. study participants of African descent

The criteria used to exclude studies were:

1. non-human/animal studies
2. review articles
3. studies reporting on adults or late-onset diabetes

### Data extraction

The data extraction was performed by two independent reviewers. The extracted data were compared and combined. The following data elements were recorded by the reviewers: 1) the last name of the first and last authors, 2) date of publication, 3) location, 4) range, mean, and median age, 5) sample size, 6) diabetic genes investigated, 7) the number of cases and controls, 8) methods used for genetic screening, 9) pathogenic gene variants found, and 10) the number of cases with the pathogenic gene variant. The data were collected and keyed into Microsoft Excel spreadsheet and analyzed using R-studio (version 4.2.1) and GraphPad Prism (version 9.5.0). Quantitative data obtained from the studies were presented using descriptive summary statistics, charts, and plots where appropriate.

### Risk of bias (quality) assessment

To avoid any form of bias, two reviewers independently synthesized the data and assessed the quality of the documents included by using the default quality assessment template in Covidence. The quality assessment was conducted at the outcome level of each study. Disagreement between the reviewers was resolved by discussion and when necessary, an expert was consulted to resolve the disagreement. The genetic tests or screening methods used by the selected studies were assessed for quality using the ACCE Model Process for Evaluating Genetic Tests.

### Evaluation of clinical significance of identified variants

The clinical significance of the variants was determined using InterVar, pathogenicity prediction tools and reports from ClinVar. InterVar is a free online bioinformatic tool and variant database which is built for variant interpretation based on the American College of Medical Genetics and Genomics and the Association for Molecular Pathology 2015 guidelines [19]. ClinVar is an aggregated database of genomic variants and their relationship with human health [20]. The following pathogenicity prediction bioinformatic tools that report meta scores were also used: BayesDel [21], MetaLR, MetaRNN, MetaSVM, and REVEL [21]. The collection of tools below with individual predictions were used to further evaluate the clinical significance of the variants: DANN, DEOGEN2, EIGEN, EIGEN PC, EVE, FATHMM, FATHMM-MKL, FATHMM-XF, LIST-S2, LRT, M-CAP [22], Mutation assessor, MutationTaster, MutPred, MVP, PrimateAI, PROVEAN, SIFT, SIFT4G, and PolyPhen (Supplementary data).

## Results

A total of 32 out of 319 records were retained after screening for data extraction (Figure 1). The earliest studies among these 32 records were conducted in 1980, while the years 2014, 2017, and 2021 recorded the highest number of studies (Figure 2A). Most early-onset diabetes genetic studies (22/32) were conducted in North African countries with the highest frequency from Egypt. There was no record from West Africa and few studies were recorded from Central and Southern Africa (Figure 2B). A review of the study designs used suggested that cross sectional and case control studies were the most preferred study designs (Figure 2C). An array of sequencing and genotyping techniques was used to determine the genetic markers associated with early-onset diabetes. These techniques include targeted and next generation sequencing, restriction fragment length polymorphism (RFLP), and human leukocyte antigens (HLA) typing (Figure 2D).

**Figure 2.**
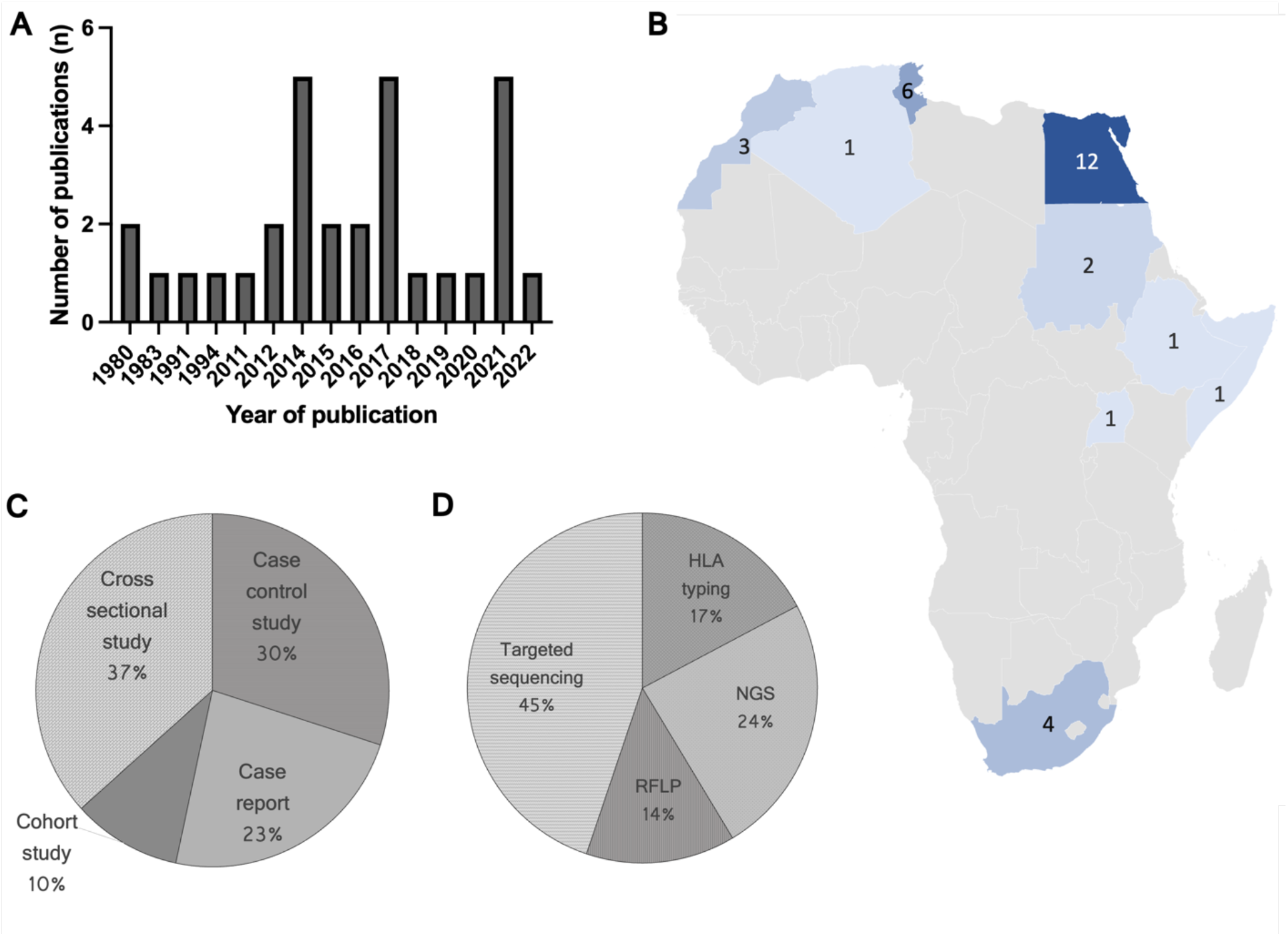
: Characteristics of publications included in this study. (A) The year of publication for the selected records. (B) Geographical representation of publications retrieved. The publications retrieved were from countries in blue shades and the numbers represent the corresponding number of publications. (C) Study design used by the various studies. (D) The molecular methods used to investigate genetics of early-onset diabetes

### Distribution of Early-onset diabetes genes in Africa

Most of the studies reported on participants with less than one year age of onset. Only 5 publications studied people with more than 18 but less than 30 years of age of onset (Figure 3A). A review of the type of diabetes revealed that most of the participants (about 600 cases) were diagnosed of type 1 diabetes (Figure 3C).

**Figure 3.**
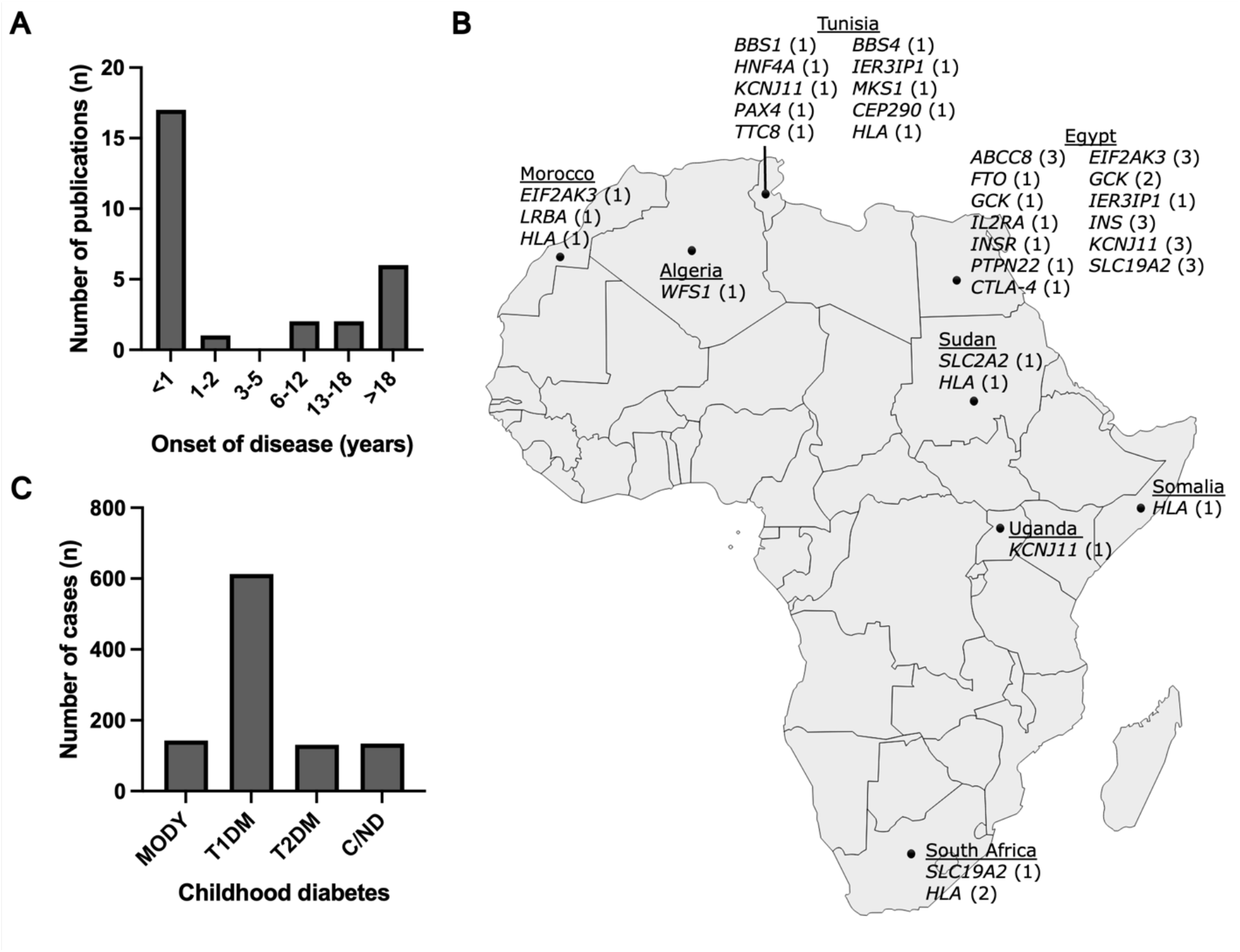
: Characteristics of study participants and distribution of identified genes. (A) Age of onset of diabetes. (B) Geographical distributions of genes associated with early-onset diabetes in Africa. The number of studies reporting on the associated genes were written in parenthesis. (C) Distribution of early-onset diabetes types diagnosed in Africa. Maturity onset diabetes of the young (MODY), type 1 diabetes (T1DM), type 2 diabetes (T2DM), unidentified childhood/neonatal diabetes (C/ND).

Analysis of the data retrieved showed that variants in 22 genes were associated with early-onset diabetes cases in Africa (Supplementary data). Six studies conducted HLA typing to determine the distribution of HLA antigens in diabetic patients. Except for HLA, *KCNJ11* was the most studied gene (5 publications) followed by *EIF2AK3* and *SLC19A2* worked on by 4 studies each (Figure 3B). Most of the associated genes were reported from Egypt and Tunisia (Figure 3B).

### Pathogenic and likely pathogenic variants identified from early-onset diabetes cases

Among the 22 early-onset diabetes associated genes, pathogenic and likely pathogenic (PLP) variants were found in 9 genes. A total of 64 variants were identified in the 22 early-onset diabetes associated genes of which 19 likely pathogenic variants were found in the 9 genes (Table 1). Seven of the PLP variants were reported on the homozygous state while 8 variants were heterozygous. The most common PLP variants was *IER3IP1*: p.(Leu78Pro) reported in 4 unrelated cases. The majority (16/19 variants) of the PLP variants were non-synonymous with a few (4/19 variants) resulting in premature termination of a protein (Table 1).

**Table 1:** A list of pathogenic and likely pathogenic variants.

| Gene | Nucleotide change | Protein change | Unrelated cases | Country | Reference |
| --- | --- | --- | --- | --- | --- |
| *ABCC8* | (NM_000352.6):c.1792C>T | p.(Arg598Ter) | 2(het) | Egypt | [23] |
| *ABCC8* | (NM_000352.6):c.970G>A | p.(Val324Met) | 1 (het) | Egypt | [24] |
| *ABCC8* | (NM_001287174.2):c.4220G>T | p.(Gly1407Val) | 1 (het) | Egypt | [24] |
| *ABCC8* | (NM_000352.6):c.3766G>A | p.(Ala1256Thr) | 1(hom) | Egypt | [24] |
| *ABCC8* | (NM_001287174.2):c.4138C>T | p.(Arg1380Cys) | 2 | Egypt | [25] |
| *ABCC8* | (NM_000352.6):c.1069G>T | p.(Val357Phe) | 1 | Egypt | [25] |
| *EIF2AK3* | (NM_004836.7):c.1958G>C | p.(Arg653Thr) | 2(hom) | Egypt | [23] |
| *EIF2AK3* | (NM_004836.7):c.1147G>T | p.(Glu383Ter) | 1(hom) | Egypt | [24] |
| *GCK* | (NM_000162.5):c.562G>A | p.(Ala188Thr) | 2(hom) | Egypt | [23] |
| *IER3IP1* | (NM_016097.5):c.233T>C | p.(Leu78Pro) | 4 | Egypt | [26] |
| *IER3IP1* | (NM_016097.5):c.62T>G | p.(Val21Gly) | 1(hom) | Tunisia | [27] |
| *INS* | (NM_001185097.2):c.265C>T | p.(Arg89Cys) | 1(hom) | Egypt | [24] |
| *KCNJ11* | (NM_000525.4):c.521C>G | p.(Ala174Gly) | 2(het) | Egypt | [23] |
| *KCNJ11* | (NM_000525.4):c.601C>T | p.(Arg201Cys) | 1(het) | Egypt | [28] |
| *KCNJ11* | (NM_000525.4):c.602G>T | p.(Arg201Leu) | 1(het) | Egypt | [29] |
| *KCNJ11* | (NM_000525.4):c.679G>A | p.(Glu227Lys) | 1 (het) | Tunisia | [30] |
| *LRBA* | (NM_006726.4):c.7042C>T | p.(Arg2348Ter) | 1 | Morocco | [31] |
| *MKS1* | (NM_017777.4):c.1423C>T | p.(Arg475Cys) | 1(het) | Tunisia | [32] |
| *SLC19A2* | (NM_006996.3):c.1160G>A | p.(Trp387Ter) | 1(hom) | Egypt | [23] |

### Network analysis

The nine genes with pathogenic variants were used to construct a protein-protein interaction network on the STRING and gene interaction network on GeneMania databases (Figure 4). The STRING database consist of known and predicted protein-protein interactions curated from experimental data, computational predictions and publications [33]. Genemania is a database for predicting gene interactions and consist of over 660 million network interactions [34]. Two proteins, ABCC8 and KCNJ11, were found to have the highest number of connections in both STRING and GeneMania interaction networks (Figure 4A and B) suggesting their in the network. Gene ontology (GO) analysis was conducted with the all the genes retrieved from the networks. Impaired glucose tolerance and decreased insulin secretion were found to be the top two GO hits obtained from the MGI Mammalian Phenotype Level4 2021 onthology (Figure 4C). Similarly, ion channels were found to be the cell components associated with the disease, hence may explain the disease mechanism and serve as targets for new drugs (Figure 4D).

**Figure 4.**
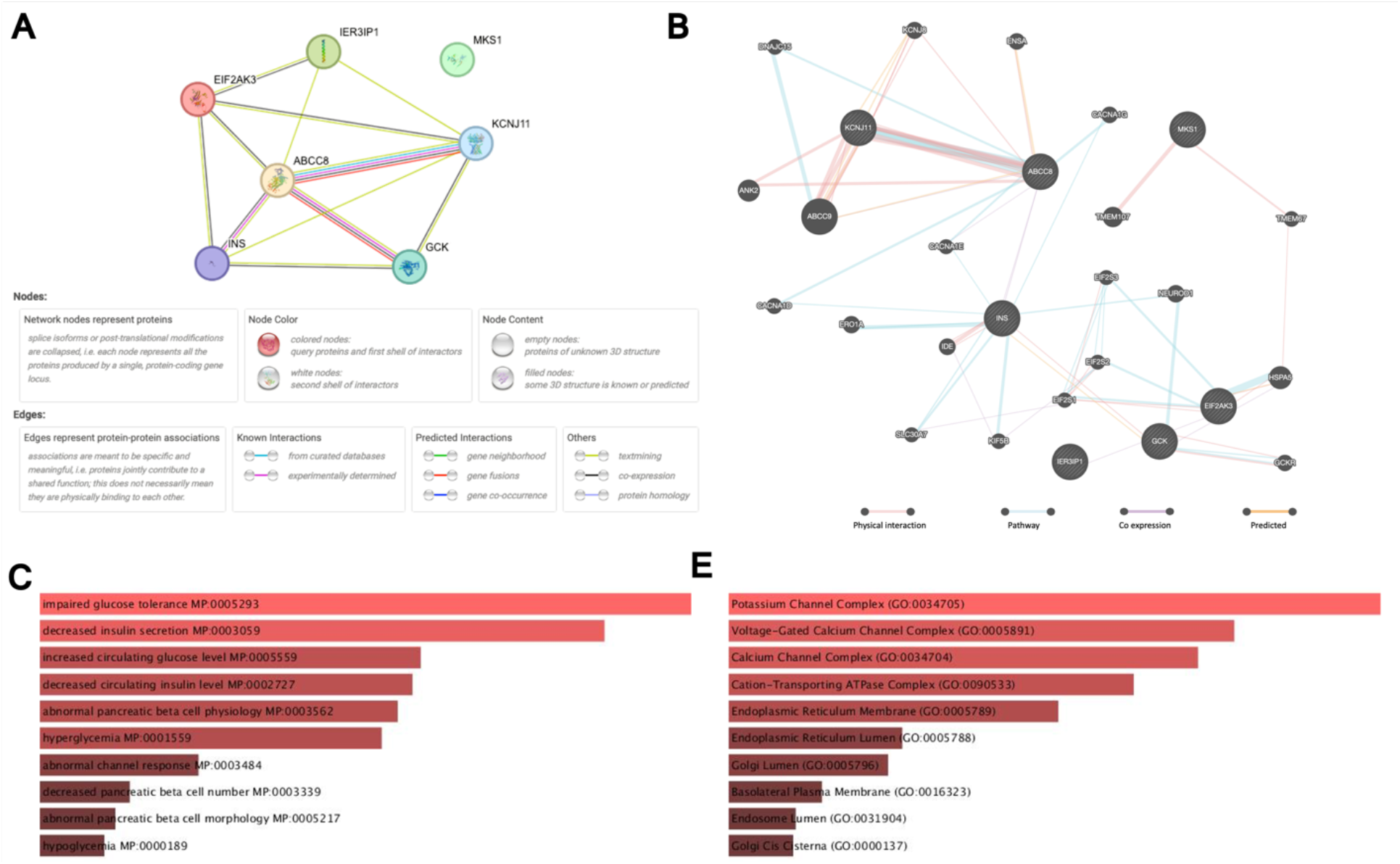
Earl-onset diabetes related interaction networks: (A) protein-protein interaction network from the STRING database. (B) Gene interaction network from GeneMania database. The size of the nodes corresponds to the number of connections. Gene ontology analysis of early onset diabetes genes showing the top hits from (C) MGI Mammalian Phenotype Level4 2021 ontology and (D) GO Cellular Component 2023. The ontologies were obtained obtained from Enrichr database [35].

## Discussion

In the past decade, there has been a rise in the global prevalence of early-onset diabetes and in countries like China the condition is considered an epidemic [36]. The increase in the global prevalence of early-onset diabetes may be explained by an increase in sedentary lifestyle with low physical activity and diet. Diabetes is a complex trait with several gene variants [1] interacting with behavioral and environmental factors to result in the condition. Furthermore, genetic variants have been associated with poor glycemic control [37]. It is important to study the gene variants associated with early-onset diabetes to improve its diagnosis and treatment. Here, we sought to discuss the gene variants associated with early-onset diabetes in Africa and the need to increase research capacity on the continent to contribute effectively to the clinical management of the condition.

In clinical practice, especially in the developed countries, molecular genetic testing for some early-onset diabetes such as MODY are available [6, 38-40]. This advancement was preceded by several years of research efforts to identify gene variants associated with the disease. There are several studies across the globe reporting on the genetics of early-onset diabetes [38, 40, 41], however, only few of these are from Africa. Within the continent, most of the retrieved records were from North Africa with only few studies from East Africa and South Africa. There was no genetic study retrieved from West Africa. Considering the multigenic nature of diabetes, NGS emerges as the promising diagnostic tool [42], however, the majority of studies from Africa used targeted sequencing approaches. Although targeted sequencing is cheaper and require less computational power to analyze the sequence data, it may be time consuming and expensive considering the number of genes involved in the pathogenesis of diabetes. Hence, there is a need to develop cost effective and population specific targeted gene panels for an effective diagnosis of the condition.

MODY is a heterogenous early-onset diabetes with 14 associated genes mainly inherited in an autosomal dominant fashion [6]. Each gene is linked to at least a sub-type of MODY: MODY1 (*HNF4A*), MODY2 (*GCK*), MODY3 (*HNF1A*), MODY4 (*IPF1*), MODY5-(*HNF1B*), MODY6 (*NEUROD1*), MODY7 (*KLF11*), MODY8 (*CEL*), MODY9 (*PAX4*), MODY10 (*INS*), MODY11 (*BLK*), MODY12 (*ABCC8*), and MODY13 (*KCNJ11*) [43, 44]. Variants in these genes may cause pancreatic beta cells dysfunction and lead to the development of early-onset non-insulin dependent diabetes [42]. Among the 14 known MODY genes, 6 of them (*ABCC8, INS, GCK, HNF4A, KCNJ11*, and *PAX4*) were reported in African patients. *ABCC8, INS*, and *GCK* were reported from Egypt [23-25], *HNF4A* and *PAX4* from Tunisia [44, 45], and *KCNJ11* from Egypt [28, 46], Tunisia [30], and Uganda [47]. Pathogenic variants were found in 4 out of the 6 genes and these are *ABCC8, GCK, INS*, and *KCNJ11*. Sulfonylurea receptor 1 (SUR1) protein is encoded by *ABCC8* (OMIM #600509) which is located on chromosome 11p15.1 and required for normal insulin secretion. A SUR proteins are a complex of potassium channels associated with familial hyperinsulinemic hypoglycemia [48]. *GCK* (OMIM #138079) located on chromosome 7p13 codes for glucokinase, an enzyme which catalyzes the phosphorylation of glucose. The insulin gene, *INS* (OMIM#) is located on chromosome 11p15.5 and responsible for insulin production which is required for glucose control in the blood. Potassium channel, inwardly rectifying, subfamily J, member 11 (*KCNJ11*, OMIM #600937) encodes ATP-sensitive potassium channels in the pancreas, neurons, and muscle cells. *KCNJ11* is located on chromosome 11p15.1 and has similarities with the SUR proteins. The identification of these genes, mostly from North Africa, has provided some information for the delineation of the molecular mechanism of the disease which may give guidance on its clinical management.

Similar to other forms of diabetes, T2DM is heterogenous, and it has some genes (*HNF4A, HNF1A, HNF1B, PAX4*, and *KCNJ11*) in common with MODY. We have identified only three genes (*KCNJ11, HNF4A* and *PAX4*) which could be classified as T2DM genes from the African population. However, these genes were not associated with early-onset T2DM. For example, mutations in *KCNJ11* gene which was previously reported as a T2DM susceptibility gene [49], were associated with infant monogenic diabetes in an Egyptian cohort [28, 46]. *HNF4A* and *PAX4* were also associated with early-onset diabetes in Tunisian patients. T1DM on the other hand did not have several associated genes, the records retrieved rather investigated the association of HLA alleles to disease susceptibility [50-52].

Our network analysis shows that *KCNJ11* and *ABCC8* are the two genes with highest interactions. Similar results of antidiabetic drugs were obtained when the two genes were queried separately on PHAROS, a druggable genome database [53]. Below is the list of the drugs that can modulate the activities of KCNJ11 and ABCC8 proteins: acetohexamide, carbutamide, chlorpropamide, diazoxide, glibenclamide, glibornuride, gliclazide, glimepiride, glipizide, gliquidone, glisoxepide, minoxidil, mitiglinide, nateglinide, pinacidil, repaglinide, tolazamide, and tolbutamide. Most of these drugs are known antidiabetic medications used in the clinics [54-56], however they can be considered for treating African patients with early onset diabetes particularly in patients who are not responding to first line drugs such as insulin and metformin.

### Limitations and perspectives

Few articles were retrieved from West African populations which shows the limited research efforts in the sub-region. Most the West African studies were focused on late-onset diabetes or the use of natural products as antidiabetics. With the limited data from West Africa, the study could not adequately report on the genetic landscape of early-onset diabetes in Africa. In addition, diabetes is highly heterogenous with several associated genes and clinical presentations. There are several classifications and sub-classifications of the disease that share similar characteristics and phenotypes. MODY which is a form of monogenic diabetes has more than 13 different sub classifications and MODY shares some of its associated genes with T2DM [44]. Secondly, most African studies used Sanger sequencing and other single gene approaches to screen for early-onset associated genetic variants [44]. These techniques are not able to solve most cases hence the genetic causes of the disease in the African population remains largely unknown. It is therefore difficult to diagnose and manage the disease in African clinics.

We have retrieved 22 genes associated with early-onset diabetes of which 9 were found to have PLP variants. These genes can be prioritized in screening African patients in resource limited settings or used to develop a targeted NGS panel which may be relatively cheaper and more effective compared to widely used single gene approach. It is worth mentioning that a diagnostic panel of the current genes may not be useful in screening sub-Saharan Africans. Africa is genetically diverse [57], and our review highlights the need for an extensive study of the sub-Saharan African populations to enrich the collection of early-onset diabetes associated genes. With this enrichment of genes, we will be able to understand the molecular mechanisms of the disease pathogenesis as well as develop population specific molecular diagnostic tools.

## Conclusion

Investigating the molecular genetics of early-onset diabetes is crucial for understating the mechanism of the disease pathogenesis, diagnosis, and the search of effective treatment options. There are however only few studies on the genetics of early-onset diabetes from Africa with majority of them from North African countries. To identify possible genetic markers, these studies mostly used single gene approaches which are cheaper but not effective in identifying new genes. In this review, we have identified some early-onset diabetic genes which can be used to develop a diagnostic panel for screening African patients. There is a need to study other African populations, especially the sub-Sharan African populations to identify early-onset diabetes genes which can be used to improve the diagnostic efficiency of an African gene panel.

## Data Availability

All data produced in the present work are contained in the manuscript

## Supplementary Materials

The genes and variants reported from Africa and prediction of their clinical significance are tabulated in the supplementary data.

## Author Contributions

Conceptualization, S.M.A.; literature search, S.M.A., J.A.M., K.S.A., J.A., and R.O.Y.; data extraction and original draft preparation, S.M.A., J.A.M., and R.O.Y.; writing-review and editing, S.M.A., J.A.M., K.S.A., J.A., and R.O.Y. All authors contributed important intellectual content presented in this manuscript. All authors have read and agreed to the final version of the manuscript.

## Declarations

### Ethical Approval

Not applicable

## Availability of data and materials

Not applicable

## References

1. Vaxillaire M, Bonnefond A, Froguel P. The lessons of early-onset monogenic diabetes for the understanding of diabetes pathogenesis. Best practice & research Clinical endocrinology & metabolism. 2012;26(2):171–87.

2. Patterson C, Guariguata L, Dahlquist G, Soltész G, Ogle G, Silink M. Diabetes in the young–a global view and worldwide estimates of numbers of children with type 1 diabetes. Diabetes research and clinical practice. 2014;103(2):161–75.

3. Group DP. Incidence and trends of childhood type 1 diabetes worldwide 1990–1999. Diabetic medicine. 2006;23(8):857–66.

4. Sanyoura M, Philipson LH, Naylor R. Monogenic diabetes in children and adolescents: recognition and treatment options. Current diabetes reports. 2018;18:1–13.

5. Nkonge KM, Nkonge DK, Nkonge TN. The epidemiology, molecular pathogenesis, diagnosis, and treatment of maturity-onset diabetes of the young (MODY). Clinical Diabetes and Endocrinology. 2020;6(1):1–10.

6. Stride A, Hattersley AT. Different genes, different diabetes: lessons from maturity-onset diabetes of the young. Annals of medicine. 2002;34(3):207–16.

7. Naylor R. Economics of genetic testing for diabetes. Current diabetes reports. 2019;19:1–7.

8. Szopa M, Matejko B, Ucieklak D, Uchman A, Hohendorff J, Mrozińska S, et al. Quality of life assessment in patients with HNF1A-MODY and GCK-MODY. Endocrine. 2019;64:246–53.

9. Peyrot M, Rubin R, Lauritzen T, Snoek F, Matthews D, Skovlund S. Psychosocial problems and barriers to improved diabetes management: results of the Cross-National Diabetes Attitudes, Wishes and Needs (DAWN) Study. Diabetic medicine. 2005;22(10):1379–85.

10. Pasquel FJ, Lansang MC, Dhatariya K, Umpierrez GE. Management of diabetes and hyperglycaemia in the hospital. The lancet Diabetes & endocrinology. 2021;9(3):174–88.

11. Collins MM, O’Sullivan T, Harkins V, Perry IJ. Quality of life and quality of care in patients with diabetes experiencing different models of care. Diabetes care. 2009;32(4):603–5.

12. Begum M, Chittleborough C, Pilkington R, Mittinty M, Lynch J, Penno M, et al. Educational outcomes among children with type 1 diabetes: whole-of-population linked-data study. Pediatric Diabetes. 2020;21(7):1353–61.

13. Urbanová J, Brunerová L, Brož J. Hidden MODY—Looking for a Needle in a Haystack. Frontiers in Endocrinology. 2018;9:355.

14. Pinelli M, Acquaviva F, Barbetti F, Caredda E, Cocozza S, Delvecchio M, et al. Identification of candidate children for maturity-onset diabetes of the young type 2 (MODY2) gene testing: a seven-item clinical flowchart (7-iF). PLoS One. 2013;8(11):e79933.

15. Lizarzaburu-Robles JC, Gomez-de-la-Torre JC, Castro-Mujica MdC, Vento F, Villanes S, Salsavilca E, et al. Atypical hyperglycemia presentation suggests considering a diagnostic of other types of diabetes: first reported GCK-MODY in Perú. Clinical Diabetes and Endocrinology. 2020;6:1–5.

16. Al-Kandari H, Al-Abdulrazzaq D, Davidsson L, Al-Mulla F. Maturity-onset diabetes of the young (MODY): a time to act. The Lancet Diabetes & Endocrinology. 2020;8(7):565–6.

17. Schiavo JH. PROSPERO: an international register of systematic review protocols. Medical reference services quarterly. 2019;38(2):171–80.

18. Babineau J. Product review: Covidence (systematic review software). Journal of the Canadian Health Libraries Association/Journal de l’Association des bibliothèques de la santé du Canada. 2014;35(2):68–71.

19. Li Q, Wang K. InterVar: clinical interpretation of genetic variants by the 2015 ACMG-AMP guidelines. The American Journal of Human Genetics. 2017;100(2):267–80.

20. Landrum MJ, Lee JM, Riley GR, Jang W, Rubinstein WS, Church DM, et al. ClinVar: public archive of relationships among sequence variation and human phenotype. Nucleic acids research. 2014;42(D1):D980–D5.

21. Tian Y, Pesaran T, Chamberlin A, Fenwick RB, Li S, Gau C-L, et al. REVEL and BayesDel outperform other in silico meta-predictors for clinical variant classification. Scientific Reports. 2019;9(1):1–6.

22. Jagadeesh KA, Wenger AM, Berger MJ, Guturu H, Stenson PD, Cooper DN, et al. M-CAP eliminates a majority of variants of uncertain significance in clinical exomes at high sensitivity. Nature genetics. 2016;48(12):1581–6.

23. Madani H, Elkaffas R, Alkholy B, Musa N, Shaalan Y, Hassan M, et al. Identification of novel variants in neonatal diabetes mellitus genes in Egyptian patients with permanent NDM. INTERNATIONAL JOURNAL OF DIABETES IN DEVELOPING COUNTRIES. 2019;39(1):53–9. doi: 10.1007/s13410-018-0658-6.

24. Abdelmeguid Y, Mowafy EW, Marzouk I, De Franco E, ElSayed S. Clinical and molecular characteristics of infantile-onset diabetes mellitus in Egypt. Ann Pediatr Endocrinol Metab. 2022. doi: 10.6065/apem.2142184.092.

25. Laimon W, El-Ziny M, El-Hawary A, Elsharkawy A, Salem NA, Aboelenin HM, et al. Genetic and clinical heterogeneity of permanent neonatal diabetes mellitus: a single tertiary centre experience. Acta Diabetol. 2021;58(12):1689–700. doi: 10.1007/s00592-021-01788-6.

26. Abdel-Salam GM, Schaffer AE, Zaki MS, Dixon-Salazar T, Mostafa IS, Afifi HH, et al. A homozygous IER3IP1 mutation causes microcephaly with simplified gyral pattern, epilepsy, and permanent neonatal diabetes syndrome (MEDS). Am J Med Genet A. 2012;158A(11):2788–96. doi: 10.1002/ajmg.a.35583.

27. Rjiba K, Soyah N, Kammoun M, Hadj Hmida I, Saad A, McElreavey K, et al. Further report of MEDS syndrome: Clinical and molecular delineation of a new Tunisian case. Eur J Med Genet. 2021;64(9):104285. doi: 10.1016/j.ejmg.2021.104285.

28. Madani HA, Fawzy N, Afif A, Abdelghaffar S, Gohar N. STUDY OF KCNJ11 GENE MUTATIONS IN ASSOCIATION WITH MONOGENIC DIABETES OF INFANCY AND RESPONSE TO SULFONYLUREA TREATMENT IN A COHORT STUDY IN EGYPT. ACTA ENDOCRINOLOGICA-BUCHAREST. 2016;12(2):157–60. doi: 10.4183/aeb.2016.157.

29. Ahmed DM, Abdel Dayem SM, Abdel Kader M, Khalifa RH, El-Lebedy DH, Kamel SA, et al. Utilizing the KCNJ11 Gene Mutations in Spotting Egyptian Patients With Permanent Neonatal Diabetes Who Can Benefit From Treatment Shift. Lab Med. 2017;48(3):225–9. doi: 10.1093/labmed/lmw067.

30. Kamoun T, Chabchoub I, Ben Ameur S, Kmiha S, Aloulou H, Cave H, et al. Transient neonatal diabetes mellitus and activating mutation in the KCNJ11 gene in two siblings. Arch Pediatr. 2017;24(5):453–6. doi: 10.1016/j.arcped.2017.02.021.

31. Johnson MB, De Franco E, Lango Allen H, Al Senani A, Elbarbary N, Siklar Z, et al. Recessively Inherited LRBA Mutations Cause Autoimmunity Presenting as Neonatal Diabetes. Diabetes. 2017;66(8):2316–22. doi: 10.2337/db17-0040.

32. Dallali H, Kheriji N, Kammoun W, Mrad M, Soltani M, Trabelsi H, et al. Multiallelic Rare Variants in BBS Genes Support an Oligogenic Ciliopathy in a Non-obese Juvenile-Onset Syndromic Diabetic Patient: A Case Report. Front Genet. 2021;12:664963. doi: 10.3389/fgene.2021.664963.

33. Szklarczyk D, Franceschini A, Kuhn M, Simonovic M, Roth A, Minguez P, et al. The STRING database in 2011: functional interaction networks of proteins, globally integrated and scored. Nucleic acids research. 2010;39(suppl_1):D561–D8.

34. Montojo J, Zuberi K, Rodriguez H, Bader GD, Morris Q. GeneMANIA: Fast gene network construction and function prediction for Cytoscape. F1000Research. 2014;3.

35. Xie Z, Bailey A, Kuleshov MV, Clarke DJ, Evangelista JE, Jenkins SL, et al. Gene set knowledge discovery with Enrichr. Current protocols. 2021;1(3):e90.

36. Pan J, Jia W. Early-onset diabetes: an epidemic in China. Frontiers of medicine. 2018;12:624–33.

37. Alfaqih MA, Al-Hawamdeh A, Amarin ZO, Khader YS, Mhedat K, Allouh MZ. Single nucleotide polymorphism in the ADIPOQ gene modifies adiponectin levels and glycemic control in type two diabetes mellitus patients. BioMed Research International. 2022;2022.

38. Shields B, Hicks S, Shepherd M, Colclough K, Hattersley AT, Ellard S. Maturity-onset diabetes of the young (MODY): how many cases are we missing? Diabetologia. 2010;53:2504–8.

39. Naylor RN, John PM, Winn AN, Carmody D, Greeley SAW, Philipson LH, et al. Cost-effectiveness of MODY genetic testing: translating genomic advances into practical health applications. Diabetes care. 2014;37(1):202–9.

40. Thurber BW, Carmody D, Tadie EC, Pastore AN, Dickens JT, Wroblewski KE, et al. Age at the time of sulfonylurea initiation influences treatment outcomes in KCNJ11-related neonatal diabetes. Diabetologia. 2015;58:1430–5.

41. Kanakatti Shankar R, Pihoker C, Dolan LM, Standiford D, Badaru A, Dabelea D, et al. Permanent neonatal diabetes mellitus: prevalence and genetic diagnosis in the SEARCH for Diabetes in Youth Study. Pediatric diabetes. 2013;14(3):174–80.

42. Firdous P, Nissar K, Ali S, Ganai BA, Shabir U, Hassan T, et al. Genetic testing of maturity-onset diabetes of the young current status and future perspectives. Frontiers in endocrinology. 2018;9:253.

43. Bonnefond A, Philippe J, Durand E, Dechaume A, Huyvaert M, Montagne L, et al. Whole-exome sequencing and high throughput genotyping identified KCNJ11 as the thirteenth MODY gene. PloS one. 2012;7(6):e37423.

44. Amara A, Chadli-Chaieb M, Chaieb L, Saad A, Gribaa M. Challenges for molecular diagnosis of familial early-onset diabetes in unexplored populations. Iran J Public Health. 2014;43(7):1011–3.

45. Amara A, Chadli-Chaieb M, Ghezaiel H, Philippe J, Brahem R, Dechaume A, et al. Familial early-onset diabetes is not a typical MODY in several Tunisian patients. Tunis Med. 2012;90(12):882–7.

46. Gohar NA, Rabie WA, Sharaf SA, Elsharkawy MM, Mira MF, Tolba AO, et al. Identification of insulin gene variants in neonatal diabetes. J Matern Fetal Neonatal Med. 2017;30(9):1035–40. doi: 10.1080/14767058.2016.1199674.

47. Nyangabyaki-Twesigye C, Muhame MR, Bahendeka S. Permanent neonatal diabetes mellitusa case report of a rare cause of diabetes mellitus in East Africa. Afr Health Sci. 2015;15(4):1339–41. doi: 10.4314/ahs.v15i4.37.

48. Glaser B, Chiu K, Anker R, Nestorowicz A, Landau H, Ben-Bassat H, et al. Familial hyperinsulinism maps to chromosome 11p14–15.1, 30 cM centromeric to the insulin gene. Nature genetics. 1994;7(2):185–8.

49. Hani E, Boutin P, Durand E, Inoue H, Permutt M, Velho G, et al. Missense mutations in the pancreatic islet beta cell inwardly rectifying K+ channel gene (KIR6. 2/BIR): a meta-analysis suggests a role in the polygenic basis of Type II diabetes mellitus in Caucasians. Diabetologia. 1998;41:1511–5.

50. Briggs BR, Jackson WP, DuToit ED, Botha MC. The histocompatibility (HLA) antigen distribution in diabetes in southern African Blacks (Xhosa). Diabetes. 1980;29(1):68–71.

51. Hammond MG, Asmal AC. HLA and insulin dependent diabetes in South African Indians. Tissue Antigens. 1980;15(3):244–8. doi: 10.1111/j.1399-0039.1980.tb00914.x.

52. Shires R, Maier G, Lustig A, Barnett P, Joffe BI, Seftel HC. HLA antigens in White and Black South African diabetics. S Afr Med J. 1983;64(28):1087–9.

53. Nguyen D-T, Mathias S, Bologa C, Brunak S, Fernandez N, Gaulton A, et al. Pharos: collating protein information to shed light on the druggable genome. Nucleic acids research. 2017;45(D1):D995–D1002.

54. Kecskemeti V, Bagi Z, Pacher P, Posa I, Kocsis E, Koltai M. New trends in the development of oral antidiabetic drugs. Current Medicinal Chemistry. 2002;9(1):53–71.

55. Seltzer HS. Drug-induced hypoglycemia: a review of 1418 cases. Endocrinology and metabolism clinics of North America. 1989;18(1):163–83.

56. Bajaj S, Kalra S. Second-generation Sulfonylureas. Drugs in Diabetes. 2021:22.

57. Campbell MC, Tishkoff SA. African genetic diversity: implications for human demographic history, modern human origins, and complex disease mapping. Annu Rev Genomics Hum Genet. 2008;9:403–33.

